# Donor whole blood DNA methylation is not a strong predictor of acute graft versus host disease in unrelated donor allogeneic haematopoietic cell transplantation

**DOI:** 10.1101/2022.03.08.22272071

**Authors:** Amy P. Webster, Simone Ecker, Ismail Moghul, Pawan Dhami, Sarah Marzi, Dirk S. Paul, Michelle Kuxhausen, Stephanie J. Lee, Stephen R. Spellman, Tao Wang, Andrew Feber, Vardhman Rakyan, Karl S. Peggs, Stephan Beck

**Affiliations:** UCL Cancer Institute, University College London, London, UK; College of Medicine and Health, The University of Exeter Medical School, Exeter, UK; NIHR Biomedical Research Centre, Guy’s Hospital London, London, UK; Blizard Institute, Barts and The London School of Medicine and Dentistry, London, UK; Cardiovascular Epidemiology Unit, Department of Public Health and Primary Care, University of Cambridge, Cambridge, UK; Center for International Blood and Marrow Transplant Research, National Marrow Donor Program, Minneapolis, MN; Center for International Blood and Marrow Transplant Research, Medical College of Wisconsin, Milwaukee, WI; Fred Hutchinson Cancer Research Center, University of Washington, Seattle, WA; Division of Biostatistics, Medical College of Wisconsin, Milwaukee, WI; The Institute of Cancer Research, London, UK; Department of Haematology, University College London, London, UK

## Abstract

Allogeneic hematopoietic cell transplantation (HCT) is used to treat many blood-based disorders and malignancies. While this is an effective treatment, it can result in serious adverse events, such as the development of acute graft-versus-host disease (aGVHD). This study aimed to develop a donor-specific epigenetic classifier that could be used in donor selection in HCT to reduce the incidence of aGVHD.

The discovery cohort of the study consisted of 288 donors from a population receiving HLA-A, -B, -C and -DRB1 matched unrelated donor HCT with T cell replete peripheral blood stem cell grafts for treatment of acute leukaemia or myelodysplastic syndromes after myeloablative conditioning. Donors were selected based on recipient aGVHD outcome; this cohort consisted of 144 cases with aGVHD grades III-IV and 144 controls with no aGVHD that survived at least 100 days post-HCT matched for sex, age, disease and GVHD prophylaxis.

Genome-wide DNA methylation was assessed using the Infinium Methylation EPIC BeadChip (Illumina), measuring CpG methylation at >850,000 sites across the genome. Following quality control, pre-processing and exploratory analyses, we applied a machine learning algorithm (Random Forest) to identify CpG sites predictive of aGVHD. Receiver operating characteristic (ROC) curve analysis of these sites resulted in a classifier with an encouraging area under the ROC curve (AUC) of 0.91.

To test this classifier, we used an independent validation cohort (n=288) selected using the same criteria as the discovery cohort. Different attempts to validate the classifier using the independent validation cohort failed with the AUC falling to 0.51. These results indicate that donor DNA methylation may not be a suitable predictor of aGVHD in an HCT setting involving unrelated donors, despite the initial promising results in the discovery cohort.

Our work highlights the importance of independent validation of machine learning classifiers, particularly when developing classifiers intended for clinical use.

## Introduction

In the past six decades, allogeneic hematopoietic cell transplantation (HCT) has become a cornerstone of treatment for haematological malignancies and is still often considered the only curative option^1^. Despite advances in the precision of HLA matching in unrelated donor selection and supportive care leading to ongoing improvements in HCT outcomes, severe graft versus host disease (GVHD) regularly occurs, increasing the risk of morbidity and mortality^2^. Acute GVHD occurs when the donor immune cells attack healthy tissue in the graft recipient, causing a range of inflammatory lesions which primarily affect the skin and digestive organs. Acute GVHD (aGVHD) typically occurs within 100 days of transplant. While the incidence has decreased in the last decade due to better HLA matching of donors, aGVHD still affects ∼30-50% of allogeneic HCT recipients^3^, making the prevention of aGVHD an important area of research.

DNA methylation is a stable modification of the DNA which can influence gene expression without altering the underlying genetic sequence. DNA methylation has an emerging role in precision medicine due to the environmental and developmental exposures it can capture. Several factors associated with the development of aGVHD are also known to influence the epigenome, including age^4,5^, sex^6^ and viral infections^7^. Despite the relative infancy of the field, DNA methylation classifiers predictive of clinical outcome are now being used in the clinic, notably in oncology to guide treatment of brain tumours^8,9^. The development of machine learning algorithms and increasing size of datasets has also allowed improvement in the development of such classifiers for early diagnosis and determining subtypes of disease^10^.

In 2015, we published a pilot study investigating DNA methylation as a potential classifier of aGVHD in HCT of HLA matched sibling pairs^11^. In that study, we assessed DNA methylation in a cohort of 85 HCT donors selected based on recipient outcome, identifying 31 DNA methylation markers associated with aGVHD severity in graft recipients. In internal cross-validation these markers showed strong predictive performance (AUC=0.98) indicating the potential utility of DNA methylation in improving donor selection in sibling HCT. The purpose of the current study was to investigate if DNA methylation is also predictive of outcome in HLA matched unrelated donor-recipient pairs, which constitute a much greater proportion of HCTs. To do this, we assessed genome-wide DNA methylation of 576 individuals recruited from the Center for International Blood and Marrow Transplant Research (CIBMTR). The scale and quality of annotation of the CIBMTR donor collection allowed us to use stringent selection criteria to minimise confounding and increase our power to detect methylation differences which were predictive of the development of aGVHD following HCT.

## Methods

### Study population

The discovery study cohort consisted of 288 HLA-A, -B, -C and -DRB1 matched, unrelated donor transplants reported to the CIBMTR that had pre-transplant donor peripheral blood samples available through the CIBMTR Research Repository. Patients received a transplant between 2002 to 2017 for acute lymphoblastic leukemia (ALL), acute myelogenous leukemia (AML) and myelodysplastic syndromes (MDS) using T-cell replete peripheral blood stem cell grafts, myeloablative conditioning and tacrolimus with methotrexate or mycophenylate mofetil based GVHD prophylaxis. The population was selected as a case-control cohort with 144 cases that developed aGVHD III-IV and controls with no aGVHD. Cases and controls were matched for sex, age, disease and GVHD prophylaxis. Donors were all self-reported as Caucasian.

The validation cohort (n=288) was selected using the same criteria. Using a previously described method^12^, power calculations for the discovery study using the EPIC array for genome-wide methylation measurement were performed with genome-wide significance set at 1×10^−6^. Sample groups of 140 donors matched to recipients with grade III-IV aGvHD, and 140 donors matched to recipients with no aGVHD, would give us 88% power to detect a methylation difference of 10% between the groups, and 100% power to detect methylation differences of 25%. Several additional samples for each group were profiled to ensure adequate power even if samples were removed during quality control.

### Samples

Genomic DNA was extracted from whole blood samples obtained from CIBMTR using the QIAamp DNA Blood Mini Kit (Qiagen) at the UCL Pathology Department (discovery study) and the UCL Genomics facility (validation study). The quality and concentration of DNA was assessed using NanoDrop and Qubit (Thermo Fisher).

### Genome-wide DNA methylation profiling

For each sample, 500ng high-quality DNA was bisulphite converted using the EZ DNA methylation kit (Zymo Research), using alternative incubation conditions recommended for Illumina methylation arrays. Methylation was subsequently analysed using the Infinium MethylationEPIC array (Illumina) measuring CpG methylation at >850,000 sites across the genome. Array preparation was performed at the UCL Genomics facility using standard operating procedures. Discovery and validation cohorts were processed independent at different timepoints, but within each cohort batches were minimised by distributing comparison groups evenly across BeadChips and position on BeadChip.

### Analysis overview

All analyses were performed in R version 3.6. Samples remaining following quality control (n=282 for discovery cohort and 288 for validation cohorts) were normalised using SWAN, then problematic probes were removed including those with a detection P value >0.01, probes with a beadcount <3 in more than 5% of samples^13^, non-cg probes, probes containing any common SNPs in dbSNP^14^ and probes mapping to the X or Y chromosomes. Singular variable decomposition (SVD)^15^ and principal components analysis (PCA) were used to assess batch effects in the data, which were subsequently adjusted for using Combat^16^. Cell composition was estimated and adjusted for using the Houseman method^17^ as implemented in ChAMP^18,19^, estimating cell proportions using the Reinius reference dataset^20^. Differentially methylated positions (DMPs) were assessed using a linear model in Limma^21^.

Machine learning analysis was performed using the random forest method^22^ as implemented in the RandomForest package. Instead of using all CpG sites as input for the RandomForest analysis, a subset of 10,000 CpG sites were selected through feature selection.

A supervised approach was used, where DMPs were identified in the discovery cohort using a linear model and the top 10,000 ranked probes were used as input for the random forest analysis. An alternative unsupervised approach was also carried out where the top 10,000 probes with the largest overall beta variance across all samples in the discovery cohort were used as input for the random forest analysis.

In both cases, the classifiers were then tested on matched probe sets from the validation cohort, and sensitivity and specificity of the classifiers were calculated.

### Data availability

The participants involved in the study had been recruited under different consents which require different levels of data access. According to consent given, the corresponding data are being made available in a three-tiered data access approach:

1. Processed data (beta matrix) for all individuals (n=570) are available from the open access ‘Gene Expression Omnibus’ under accession number GSE196696. To reduce the chance of reidentification, all non-cg probes, including SNP targeting rs probes have been removed. The data are provided in both raw (unnormalized) and SWAN normalised formats.
2. Raw data (IDAT files) are available for individuals with appropriate consent (n=403 in total) from the controlled access ‘European Genome-Phenome Archive’ under accession number EGAS00001006033.
3. Raw data (IDAT files) and associated phenotype information are available for all individuals included in this study (n=570) directly from CIBMTR. Data are available under controlled access release upon reasonable request and execution of a data use agreement. Requests should be submitted to CIBMTR at and include the study reference IB17-04.

## Results

### Study Design

Unrelated donor-recipient pairs undergoing HCT were selected from the CIBMTR Research Repository, based on the aGVHD outcomes in recipients (Figure 1). Blood-based DNA methylation from donors was assessed using the Illumina EPIC arrays. Methylation differences were assessed, and random forest analysis was used to test for the presence of a classifier of aGVHD outcome.

**Figure 1:**
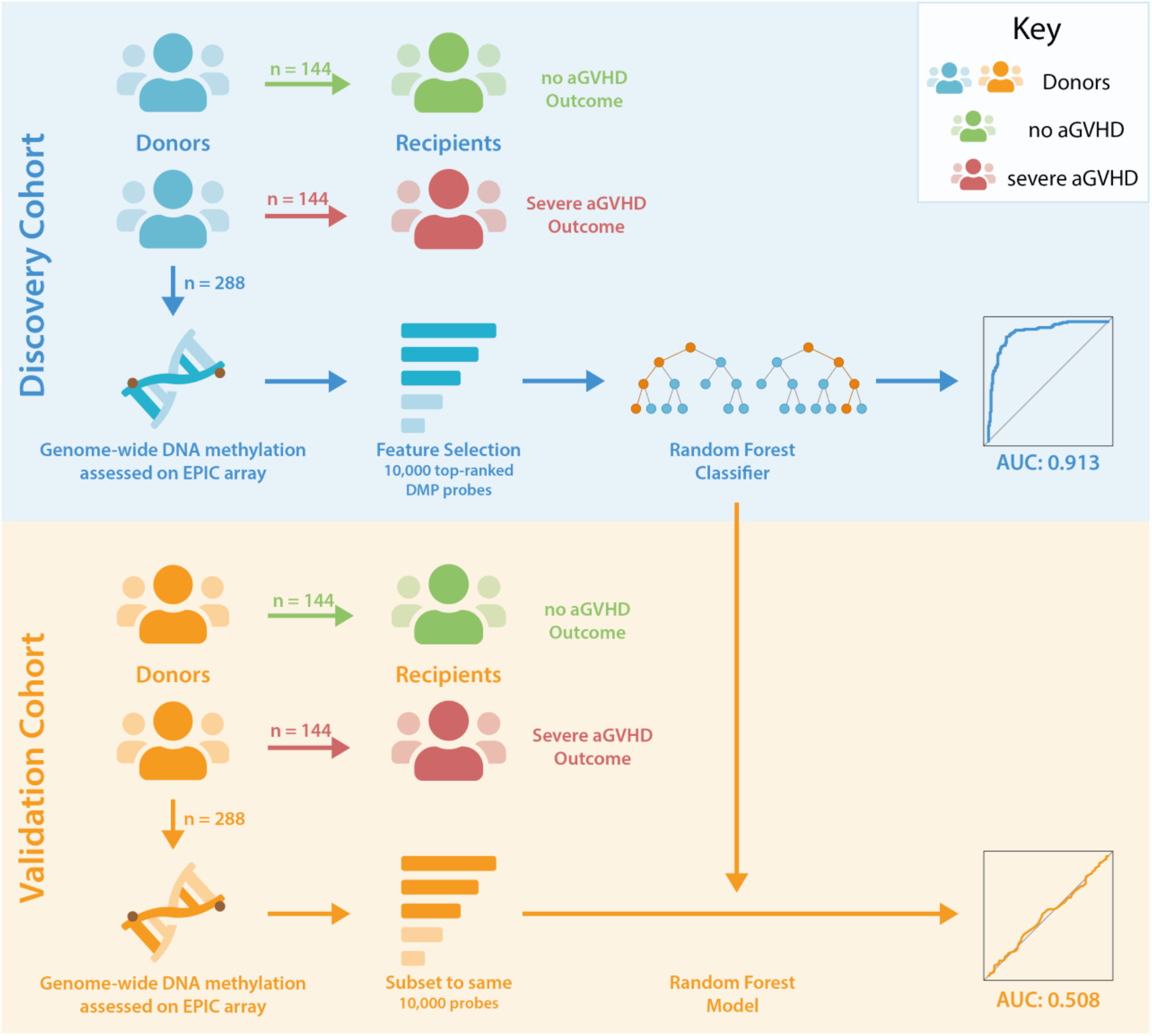
Study Design. Unrelated donor-recipient pairs were selected based on the outcome of recipients following HCT. DNA methylation levels were assessed in donors associated with no (Grade 0) or severe (Grades 3-4) aGVHD in recipients. Donor-recipient pairs were HLA matched, and comparison groups were matched for sex, age, disease and GVHD prophylaxis. Feature selection reduced the number of probes in the discovery dataset to 10,000 for input to random forest analyses, and this classifier was subsequently tested in the validation cohort following pre-processing of data and refinement to the same set of probes.

### Study population

Unrelated donor-recipient pairs were selected by CIBMTR using stringent criteria as described in methods, resulting in 282 individuals in the discovery cohort following initial data quality control, and 288 individuals in the validation cohort. The resulting cohorts were well matched for characteristics that can influence DNA methylation profile, including age and sex (as shown in Tables 1 and 2).

**Table 1:** Discovery and validation cohort characteristics. Characteristics of adult patients undergoing first allogeneic PB HCT for acute leukemia or MDS from an 8/8 HLA-matched unrelated donor between 2000-2016 with available donor blood samples, as reported to the CIBMTR. Restricted to Caucasian donors, myeloablative preparative regimens, **no ATG/Campath** and patients surviving >100 days with no aGVHD or those that developed grades III-IV aGVHD at any time post-HCT. Donors were matched between comparison groups based on sex and age by decade.

|  | Discovery cohort |  |  | Validation cohort |  |  |
| --- | --- | --- | --- | --- | --- | --- |
|  | No<br>aGVHD<br>+<br>100-day<br>survival | Grades<br>III-IV<br>aGVHD |  | No<br>aGVHD +<br>100-day<br>survival | Grades<br>III-IV<br>aGVHD |  |
| Variable | N (%) | N (%) | p-value<sup>a</sup> | N (%) | N (%) | p-value<sup>a</sup> |
| Number of Recipients | 141 | 141 |  | 144 | 144 |  |
| Disease |  |  | 0.339 |  |  | 0.020 |
| AML | 85 (60) | 73 (52) |  | 100 (69) | 77 (53) |  |
| ALL | 24 (17) | 31 (22) |  | 19 (13) | 27 (19) |  |
| MDS | 32 (23) | 37 (26) |  | 25 (17) | 40 (28) |  |
| Recipient Age |  |  | 0.464 |  |  | 1.000 |
| 18-29 | 28 (20) | 28 (20) |  | 18 (13) | 18 (13) |  |
| 30-39 | 26 (18) | 18 (13) |  | 24 (17) | 24 (17) |  |
| 40-49 | 36 (26) | 33 (23) |  | 34 (24) | 33 (23) |  |
| 50-59 | 33 (23) | 38 (27) |  | 39 (27) | 40 (28) |  |
| 60-69 | 15 (11) | 23 (16) |  | 28 (19) | 28 (19) |  |
| 70+ | 3 (2) | 1 (1) |  | 1 (1) | 1 (1) |  |
| Median (Range) | 45 (19-76) | 47 (18-72) | 0.541 | 49 (20-75) | 50 (19-71) | 0.998 |
| Recipient Sex |  |  | 0.716 |  |  | 1.000 |
| Male | 82 (58) | 85 (60) |  | 85 (59) | 85 (59) |  |
| Female | 59 (42) | 56 (40) |  | 59 (41) | 59 (41) |  |
| Recipient Race/Ethnicity |  |  | 0.113 |  |  | 1.000 |
| Caucasian | 134 (96) | 131 (94) |  | 144 (100) | 144 (100) |  |
| African American | 2 (1) | 1 (1) |  | 0 | 0 |  |
| Native American | 2 (1) | 0 |  | 0 | 0 |  |
| Caucasian, Hispanic | 2 (1) | 8 (6) |  | 0 | 0 |  |
| Unknown | 1 (N/A) | 1 (N/A) |  | 0 | 0 |  |
| Recipient ABO Type |  |  | 0.767 |  |  | 0.242 |
| A | 54 (45) | 52 (42) |  | 14 (40) | 24 (53) |  |
| B | 13 (11) | 15 (12) |  | 7 (20) | 3 (7) |  |
| AB | 3 (3) | 6 (5) |  | 4 (11) | 3 (7) |  |
| O | 50 (42) | 50 (41) |  | 10 (29) | 15 (33) |  |
| Unknown | 21 (N/A) | 18 (N/A) |  | 109 (N/A) | 99 (N/A) |  |
| Rh Factor |  |  | 0.788 |  |  | 0.167 |
| Positive | 105 (88) | 109 (89) |  | 33 (94) | 38 (84) |  |
| Negative | 15 (13) | 14 (11) |  | 2 (6) | 7 (16) |  |
| Unknown | 21 (N/A) | 18 (N/A) |  | 109 (N/A) | 99 (N/A) |  |
| Blood Type |  |  | 0.985 |  |  | 0.277 |
| A + | 46 (38) | 46 (37) |  | 14 (40) | 20 (44) |  |
| B + | 11 (9) | 13 (11) |  | 6 (17) | 3 (7) |  |
| AB + | 2 (2) | 4 (3) |  | 4 (11) | 2 (4) |  |
| O + | 46 (38) | 46 (37) |  | 9 (26) | 13 (29) |  |
| A - | 8 (7) | 6 (5) |  | 0 | 4 (9) |  |
| B - | 2 (2) | 2 (2) |  | 1 (3) | 0 |  |
| AB - | 1 (1) | 2 (2) |  | 0 | 1 (2) |  |
| O - | 4 (3) | 4 (3) |  | 1 (3) | 2 (4) |  |
| Unknown | 21 (N/A) | 18 (N/A) |  | 109 (N/A) | 99 (N/A) |  |
| Recipient CMV Status |  |  | 0.721 |  |  | 0.281 |
| Negative | 65 (46) | 65 (46) |  | 59 (41) | 69 (48) |  |
| Positive | 74 (52) | 74 (52) |  | 85 (59) | 74 (51) |  |
| Inconclusive | 1 (1) | 2 (1) |  | 0 | 1 (1) |  |
| Not tested | 1 (1) | 0 |  | 0 | 0 |  |
| Donor Age |  |  | 0.090 |  |  | 0.792 |
| 18-29 | 90 (64) | 76 (54) |  | 105 (73) | 103 (72) |  |
| 30-39 | 51 (36) | 65 (46) |  | 39 (27) | 41 (28) |  |
| Median (Range) | 26 (19-40) | 29 (19-40) | 0.003 | 26 (19-39) | 27 (19-39) | 0.076 |
| Donor Sex |  |  | 0.585 |  |  | 0.063 |
| Male | 103 (73) | 107 (76) |  | 98 (68) | 112 (78) |  |
| Female | 38 (27) | 34 (24) |  | 46 (32) | 32 (22) |  |
| Donor Race/Ethnicity |  |  | 1.000 |  |  | 1.000 |
| Caucasian | 141 (100) | 141 (100) |  | 144 (100) | 144 (100) |  |
| Donor ABO Type |  |  | 0.081 |  |  | 0.498 |
| A | 69 (49) | 51 (36) |  | 65 (45) | 60 (42) |  |
| B | 15 (11) | 14 (10) |  | 13 (9) | 8 (6) |  |
| AB | 6 (4) | 4 (3) |  | 6 (4) | 9 (6) |  |
| O | 51 (36) | 72 (51) |  | 60 (42) | 67 (47) |  |
| Donor CMV Status |  |  | 0.875 |  |  | 0.090 |
| Negative | 104 (74) | 105 (74) |  | 90 (63) | 104 (72) |  |
| Positive | 33 (23) | 32 (23) |  | 53 (37) | 38 (26) |  |
| Previously reported reactive | 1 (1) | 0 |  | 1 (1) | 0 |  |
| Not tested | 1 (1) | 1 (1) |  | 0 | 2 (1) |  |
| Unknown | 2 (1) | 3 (2) |  | 0 | 0 |  |
| Donor-Recipient Sex Match |  |  | 0.818 |  |  | 0.072 |
| Male-Male | 62 (44) | 69 (49) |  | 69 (48) | 69 (48) |  |
| Male-Female | 41 (29) | 38 (27) |  | 29 (20) | 43 (30) |  |
| Female-Male | 20 (14) | 16 (11) |  | 16 (11) | 16 (11) |  |
| Female-Female | 18 (13) | 18 (13) |  | 30 (21) | 16 (11) |  |
| Recipient Age at Diagnosis |  |  | 0.452 |  |  | 0.878 |
| < 10 | 0 | 1 (1) |  | 0 | 0 |  |
| 10-17 | 1 (1) | 4 (3) |  | 1 (1) | 0 |  |
| 18-29 | 29 (21) | 27 (19) |  | 20 (14) | 19 (13) |  |
| 30-39 | 29 (21) | 18 (13) |  | 23 (16) | 27 (19) |  |
| 40-49 | 31 (22) | 34 (24) |  | 34 (24) | 34 (24) |  |
| 50-59 | 36 (26) | 38 (27) |  | 39 (27) | 37 (26) |  |
| 60-69 | 13 (9) | 18 (13) |  | 26 (18) | 27 (19) |  |
| 70+ | 2 (1) | 1 (1) |  | 1 (1) | 0 |  |
| Median (Range) | 44 (16-75) | 46 (11-72) | 0.450 | 48 (17-75) | 49 (18-70) | 0.867 |
| MDS Disease Status |  |  | 0.465 |  |  | 0.941 |
| Early | 14 (44) | 13 (35) |  | 2 (8) | 3 (8) |  |
| Advanced | 18 (56) | 24 (65) |  | 23 (92) | 37 (93) |  |
| AML/ALL Disease Status |  |  | 0.137 |  |  | 0.009 |
| Early | 51 (47) | 43 (40) |  | 67 (56) | 67 (64) |  |
| Intermediate | 30 (28) | 21 (20) |  | 32 (27) | 11 (11) |  |
| Advanced | 23 (21) | 28 (27) |  | 20 (17) | 24 (23) |  |
| Unknown | 5 (5) | 12 (12) |  | 0 | 2 (2) |  |
| Conditioning Regimen |  |  | 0.114 |  |  | <0.001 |
| Bu + Cy | 49 (35) | 56 (40) |  | 53 (37) | 54 (38) |  |
| Bu + Mel | 1 (1) | 3 (2) |  | 0 | 0 |  |
| Bu + Flud | 38 (27) | 26 (18) |  | 61 (42) | 32 (22) |  |
| Mel + Flud | 0 | 1 (1) |  | 7 (5) | 10 (7) |  |
| Cy Alone | 43 (30) | 35 (24) |  | 22 (16) | 45(31) |  |
| Others | 10 (7) | 20 (14) |  | 1 (1) | 3 (2) |  |
| TBI Usage |  |  | 0.806 |  |  | 0.001 |
| Yes | 53 (38) | 55 (39) |  | 23 (16) | 48 (33) |  |
| No | 88 (62) | 86 (61) |  | 114 (79) | 86 (60) |  |
| Unknown | 0 | 0 |  | 7 (5) | 10 (7) |  |
| GvHD prophylaxis |  |  | 0.090 |  |  | 0.715 |
| Tac + MMF ± others | 24 (17) | 32 (23) |  | 16 (11) | 18 (13) |  |
| Tac + MTX ± others | 100 (71) | 82 (58) |  | 128 (89) | 126 (88) |  |
| CSA + MMF ± others | 4 (3) | 11 (8) |  | 0 | 0 |  |
| CSA + MTX ± others | 13 (9) | 16 (11) |  | 0 | 0 |  |
| Use of ATG or Campath |  |  | 1.000 |  |  | 1.000 |

| No ATG or<br>CAMPATH | 141<br>(100) | 141 (100) |  | 144 (100) | 144 (100) |  |
| --- | --- | --- | --- | --- | --- | --- |
| Year of<br>Transplant |  |  | 0.585 |  |  | 0.173 |
| 2002 | 0 | 1 (1) |  | 0 | 1 (1) |  |
| 2003 | 5 (4) | 4 (3) |  | 0 | 0 |  |
| 2004 | 9 (6) | 12 (9) |  | 3 (2) | 4 (3) |  |
| 2005 | 10 (7) | 13 (9) |  | 1 (1) | 6 (4) |  |
| 2006 | 8 (6) | 12 (9) |  | 1 (1) | 2 (1) |  |
| 2007 | 13 (9) | 17 (12) |  | 6 (4) | 2 (1) |  |
| 2008 | 17 (12) | 12 (9) |  | 10 (7) | 10 (7) |  |
| 2009 | 13 (9) | 19 (13) |  | 12 (8) | 14 (10) |  |
| 2010 | 23 (16) | 11 (8) |  | 13 (9) | 10 (7) |  |
| 2011 | 5 (4) | 6 (4) |  | 13 (9) | 22 (15) |  |
| 2012 | 7 (5) | 4 (3) |  | 25 (17) | 14 (10) |  |
| 2013 | 10 (7) | 13 (9) |  | 19 (13) | 19 (13) |  |
| 2014 | 9 (6) | 9 (6) |  | 21 (15) | 14 (10) |  |
| 2015 | 11 (8) | 8 (6) |  | 13 (9) | 15 (10) |  |
| 2016 | 1 (1) | 0 |  | 5 (3) | 11 (8) |  |
| 2017 | 0 | 0 |  | 2 (1) | 0 |  |

<sup>a</sup> The Pearson chi-square test was used for comparing discrete variables; the Kruskal-Wallis test was used for comparing continuous variables.

The discovery cohort was well matched for disease, with no significant difference in proportion of AML, ALL and MDS between comparison groups (p=0.339). Median recipient ages for the no/severe aGVHD groups were 45 (range 19-76) and 47 (range 18-72), respectively. There was no significant difference for recipient sex (p=0.716) or ethnicity (p=0.113) across comparison groups. Donors were well matched across comparison groups for sex (p=0.585), however, there was a difference in median age (p=0.003), though this was not apparent when individuals were stratified into age brackets (p=0.090). There were no significant differences across comparison groups for donor/recipient ABO type, blood type, Rh factor, CMV status or sex match.

The validation cohort had a significant difference in proportions of these diseases across comparison groups (p=0.02). The median recipient ages for the no/severe aGVHD groups in the validation cohort was 49 (range 20-75) and 50 (range 19-71). There was no significant difference in the recipient age distribution across comparison groups (p=0.998). There was no difference in recipient sex across the comparison groups (41% female recipients, p=1.0). Donors were well matched across comparison groups for sex (p=0.063) and median age (p=0.076). There were no significant differences in ethnicity, donor/recipient ABO type, blood type, Rh factor, CMV status or sex match across groups. There were differences in conditioning regimen across comparison groups (p<0.001).

### Data exploration and pre-processing

Following sample removal, quality control plots showed that the 282 individuals remaining in the discovery dataset and 288 individuals remaining in the validation dataset had very high quality methylation profiles (Supplementary Figure 1). Following probe filtering, 661,114 probes remained in the discovery dataset. Singular Value Decomposition (SVD) and principal components analysis (PCA) indicated that estimated ‘cell composition’, ‘Slide/BeadChip’ and ‘Array’ batch effects were having the largest impact on the data (Supplementary Figures 2-3), which were subsequently adjusted for using ChAMP cell composition correction and ComBat adjustment respectively. Cell type proportions were estimated for each group using the DNA methylation profiles and were found to be well balanced in each cohort with no significant difference between groups (Supplementary Table 1).

### Differential methylation analysis

No CpG sites passed a false discovery rate adjusted p-value significance threshold of 0.05 during DMP analysis when comparing the ‘no aGVHD’ group to the ‘severe aGVHD’ group. As the main batch and confounding effects of slide, array and cell composition had been previously adjusted in the dataset, no additional covariates were included during linear regression.

### Classifier generation from discovery data

Random forest analysis was performed on two sets of probes; the unsupervised analysis using the top-ranked 10,000 most variable probes, which all had a beta variance of >33% across all samples. The supervised analysis used the top 10,000 probes resulting from the linear model DMP analysis, though none passed statistical significance these were considered sites with putative methylation differences. Random forest analysis was run with 500 trees, with 100 variables tested at each split for both analysis approaches.

The high variability classifier showed very poor performance, with an out-of-bag (OOB) estimate of error rate of 45.39% and area under the curve (AUC) of 0.516 during internal cross-validation of the discovery dataset (Supplementary Figure 4). The differential methylation dataset produced an initially promising classifier with an OOB estimate of error rate of 14.89% and an AUC of 0.913 (Figure 2).

**Figure 2:**
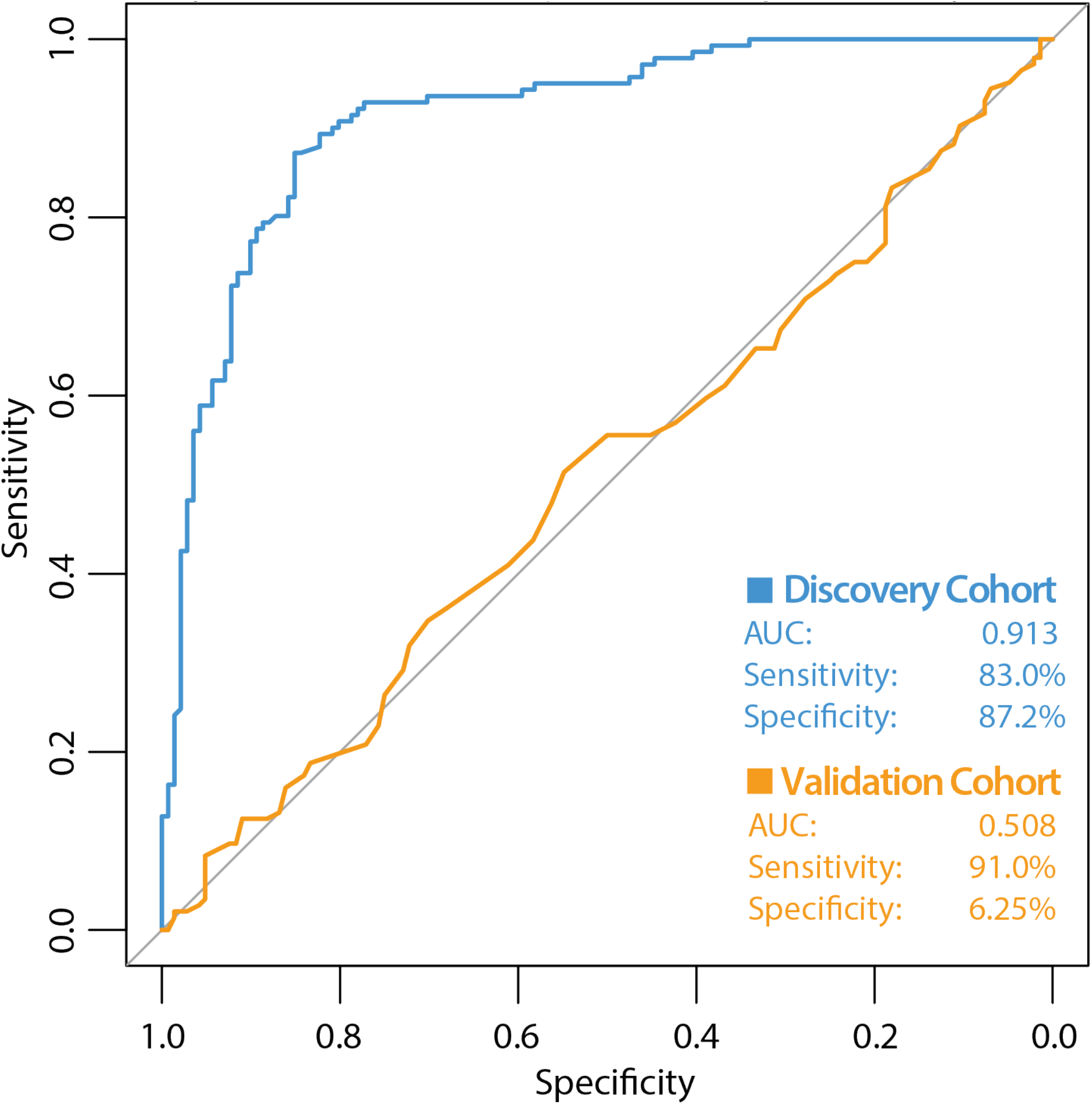
ROC curve of classifier performance of the unsupervised Random Forest Classifier. The figure shows the performance of the differential methylation (supervised approach) classifier which used the top 10,000 most differentially methylated CpG sites as input, during internal cross validation on the training dataset (blue line). The performance of the differential methylation classifier on the independent validation cohort is indicated by the orange line, which had an AUC of 0.508, a sensitivity of 90.97% with a very poor specificity of 6.25%. While initially this differential methylation-based classifier appeared encouraging with the discovery cohort, the classifier did not perform well during validation analyses.

During validation analysis, the matched CpG sites used as input to the original random forest training analysis were extracted from the validation dataset as all probes present in training analyses are required as input for validation. Validation analyses indicated that the differential methylation classifier had a sensitivity of 90.97% but a specificity of just 6.25%, and an AUC of 0.508. This is driven by an over-prediction of the ‘severe aGVHD’ group in the independent validation cohort, resulting in many false positive predictions. The unsupervised differential variability classifier also had an extremely poor performance in the validation cohort, with a sensitivity of just 50%, a specificity of 51.39% and an AUC of 0.523. As such, neither of these approaches yielded a useful classifier.

## Discussion

Recently developed predictors of aGVHD using clinical variables have had modest success with an AUC of ∼ 0.6^23^, however this indicated that biological markers of gene expression, such as epigenetic markers, could provide additional insight to improve prediction of aGVHD. This was also supported by the recent finding that hypermethylation of the TP53 gene in HCT recipients was found to correlate with relapse of myelodysplastic syndromes following transplantation, indicating recipient-based DNA methylation could be predictive of outcomes during HCT^24^. As DNA methylation levels reflect both the underlying genetic sequence and factors known to be associated with aGVHD development (such as donor age, sex and cytomegalovirus serostatus), we hypothesised they would be a strong candidate for classifier identification. Our initial study focused on sibling donor-recipient pairs, in which a DNA methylation classifier of aGVHD development was identified in the blood of donors^11^. In the current study, we tested if DNA methylation as measured by EPIC arrays is also predictive of aGVHD in unrelated donor-recipient pairs and found that it is not.

There are several potential technical and biological reasons that a robust classifier of aGVHD was not identified in this study. Firstly, while the study performed was shown to have power to detect larger methylation differences of >10%, the relatively small sample size of the discovery cohort (n=280) and validation cohort (n=288) may have limited our ability to detect more subtle methylation differences. In the future, larger scale studies may provide increased power to detect such differences.

Secondly, the tissue we investigated was peripheral blood of donors which was intended to act as a surrogate tissue reflecting outcome. DNA methylation profiles are known to be highly cell type specific^25^, and while blood based DNA methylation may reflect certain exposures and factors associated with aGVHD development, it is possible that a specific cellular subtype which is not present in the whole blood of donors is responsible for the development of aGVHD and as such would not be reflected in the methylation profile.

Another possibility is that the specific cell type which is causing aGVHD could be present in whole blood, but in small proportions, making the signal significantly diluted by other more prominent cell types. Indeed, in the current analysis, cell composition was the biggest driver of variation in the data, and though this was balanced overall between the comparison groups and adjusted for in the data analysis, it could have been a confounding factor in the study, or subtle methylation effects could have been lost during adjustment. In the future, methylation analysis of individual cell types isolated from stem cell grafts may provide more insight into DNA methylation differences driving the development of aGVHD. While this approach would provide a more refined methylation measurement, it would be a significantly less practical approach for a clinical test, limiting the utility for optimising donor selection as usually these cells would only be collected once a donor is committed.

A classifier of aGVHD development was identified in our previously published work, which investigated donor DNA methylation from sibling HCT. A potential reason a similar biomarker was not identified in this cohort is that it could have been specific to sibling transplants, which generally have a lower incidence of aGVHD which may be driven more by extrinsic factors which influence DNA methylation, while aGVHD following HCT from an unrelated donor may be driven more by genetic factors. There may also be an issue of ‘epigenetic compatibility’, with donors and recipients varying in epigenetic profile inciting the initiation of aGVHD in certain individuals, without this being driven by a specifically differentially methylated gene or pathway. This would explain why a classifier was not identified in the current study, as the epigenetic marks conferring risk of aGVHD would be different for each individual. In the future, studies investigating the DNA methylation of both donors and recipients during HCT could provide more insight into this possibility.

When considering the clinical context of the development of aGVHD, it is likely the end result of a complicated clinical setting with multiple donor and recipient factors affecting the outcome. If the epigenetic pattern was highly predictive, it might infer that the occurrence of severe aGVHD is pre-ordained just by donor factors, which seems biologically unlikely.

On a technical level, this study has also demonstrated the importance of careful development and testing of analysis pipelines for methylation studies, in particular when applying complex machine learning methods to datasets. Our initial findings indicated a robust classifier might be present within the dataset, a finding which was amplified when data was pre-processed as a single batch with subsequent splitting of the dataset and internal cross validation. While our validation dataset was of exceptionally high quality and donors included were matched to a very high degree with the discovery cohort, the classifier was not validated even with extensive optimisation and testing of alternate pipeline settings. This demonstrates that even with the identification of a promising and robust classifier in a well-designed study, independent validation is critical^26^, and such validation datasets need to be generated completely independently with unique individuals and pre-processed separately to the training/discovery cohort. This also better mimics the experimental realities of clinical classifier use, making any findings that do stand up to the validation process more robust and clinically useful.

## Conclusions

In this study, we performed the definitive investigation of donor-derived blood-based DNA methylation as a classifier of aGVHD outcome in HCT and found that donor DNA methylation as assessed by methylation arrays is not a strong candidate for prediction of aGVHD. It is possible that other methylation signals exist which might improve our understanding of the development of aGVHD in these cohorts, which we plan to investigate in the future. We have also highlighted the importance of study design and well-designed independent validation of methylation differences especially when applying machine learning approaches.

## Data Availability

The participants involved in the study had been recruited under different consents which require different levels of data access. According to consent given, the corresponding data are being made available in a three-tiered data access approach:
1. Processed data (beta matrix) for all individuals (n=570) are available from the open access 'Gene Expression Omnibus' under accession number GSE196696. To reduce the chance of reidentification, all non-cg probes, including SNP targeting rs probes have been removed. The data are provided in both raw (unnormalized) and SWAN normalised formats.
2. Raw data (IDAT files) are available for individuals with appropriate consent (n=403 in total) from the controlled access 'European Genome-Phenome Archive' under accession number EGAxxxxx.
3. Raw data (IDAT files) and associated phenotype information are available for all individuals included in this study (n=570) directly from CIBMTR. Data are available under controlled access release upon reasonable request and execution of a data use agreement. Requests should be submitted to CIBMTR at and include the study reference IB17-04.

https://www.ncbi.nlm.nih.gov/geo/

https://ega-archive.org

## Acknowledgements

The authors would like to thank UCL Genomics and UCL Pathology for their support with DNA extraction and array preparation.

This project was funded by the National Institute for Health Research (NIHR) Blood & Transplant Research Unit (BTRU) (NIHR-BTRU-2014-10074). The views expressed are those of the authors and not necessarily those of the NIHR or the Department of Health and Social Care. The CIBMTR is supported primarily by Public Health Service U24CA076518 from the National Cancer Institute (NCI), the National Heart, Lung and Blood Institute (NHLBI) and the National Institute of Allergy and Infectious Diseases (NIAID); HHSH250201700006C from the Health Resources and Services Administration (HRSA); and N00014-20-1-2705 and N00014-20-1-2832 from the Office of Naval Research; Support is also provided by Be the Match Foundation, the Medical College of Wisconsin, and the National Marrow Donor Program.

Finally, the authors would like to sincerely thank all donors, patients and their families who contributed to CIBMTR cohort.

