## Supplementary Figure 1 for "Donor whole blood DNA methylation is not a strong predictor of acute graft versus host disease in unrelated donor allogeneic haematopoietic cell transplantation"

**Supplementary Figure 1: Quality control beta distribution plots.** The beta distributions for the raw (unnormalized) beta distributions of discovery and validation datasets show that the array performed very well and the data was of exceptionally high quality.

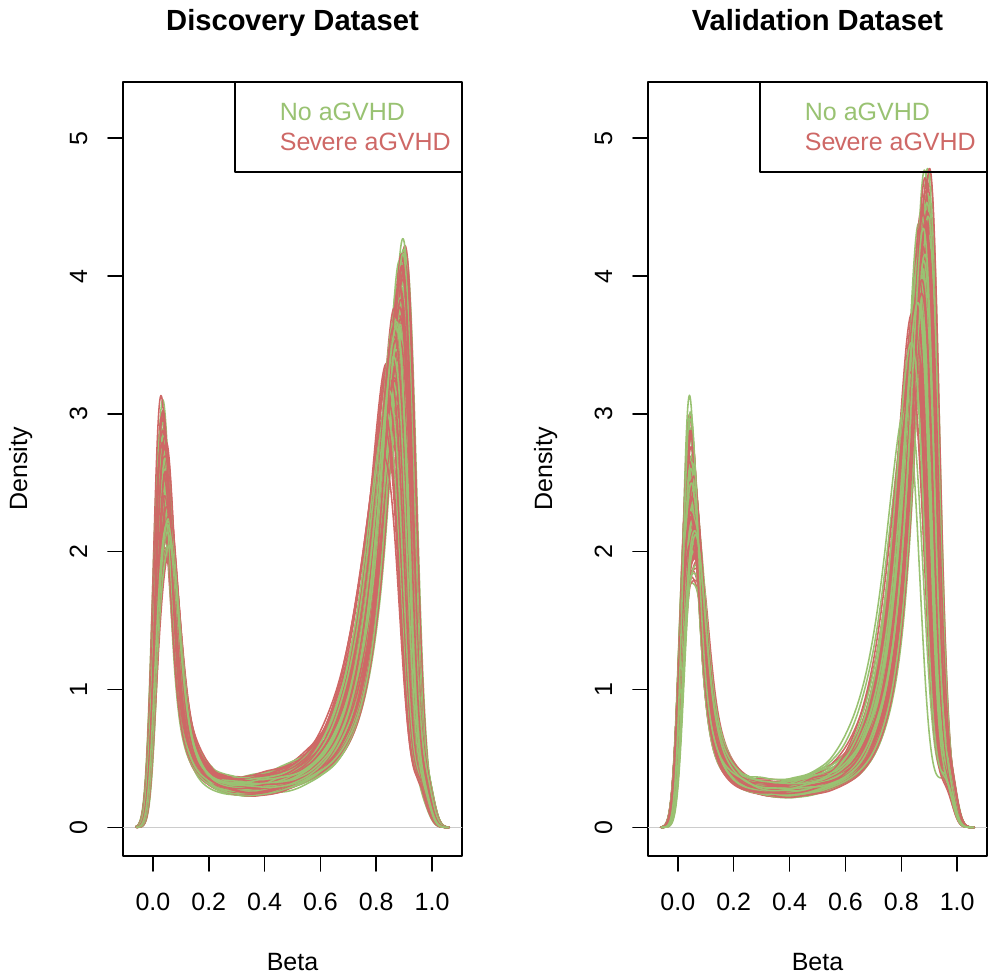

**Supplementary Figure 2: SVD plot indicating batch effects in the discovery dataset.** Singular value decomposition (SVD) plot shown for the SWAN normalised dataset with problematic probes removed to minimise the effect of the confounding of donor sex on the dataset. This indicates that top principal components are strongly associated with BeadChip, cell composition and Array (ie, position on BeadChip). These factors were subsequently corrected for during data pre-processing.

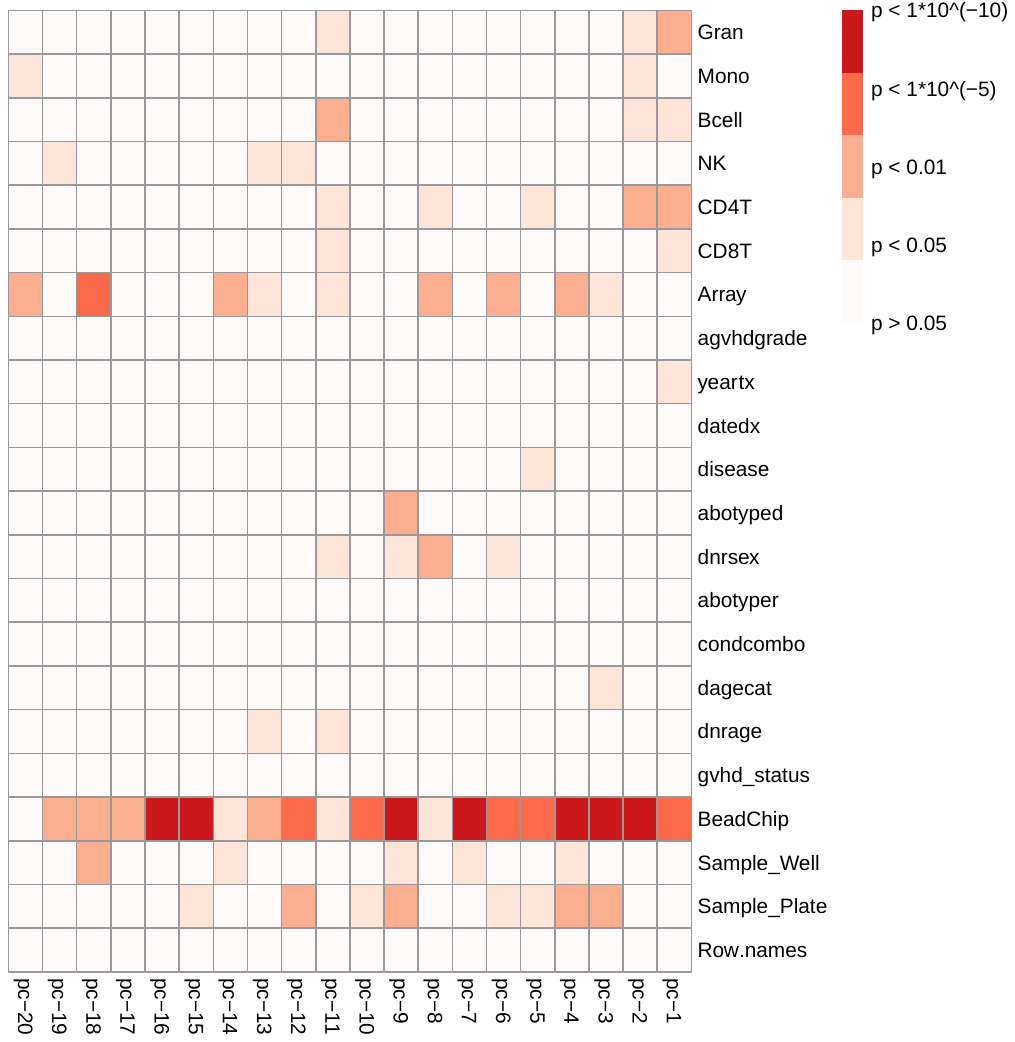

**Supplementary Figure 3: PCA plot of top two principal components and the distribution of ‘severe aGVHD’ and ‘no aGVHD’ comparison groups.** Principal Components Analysis (PCA) showed that in the top two principal components, which explain 12.7% and 8.96% of the variation in the dataset respectively, there was no correlation with aGVHD status.

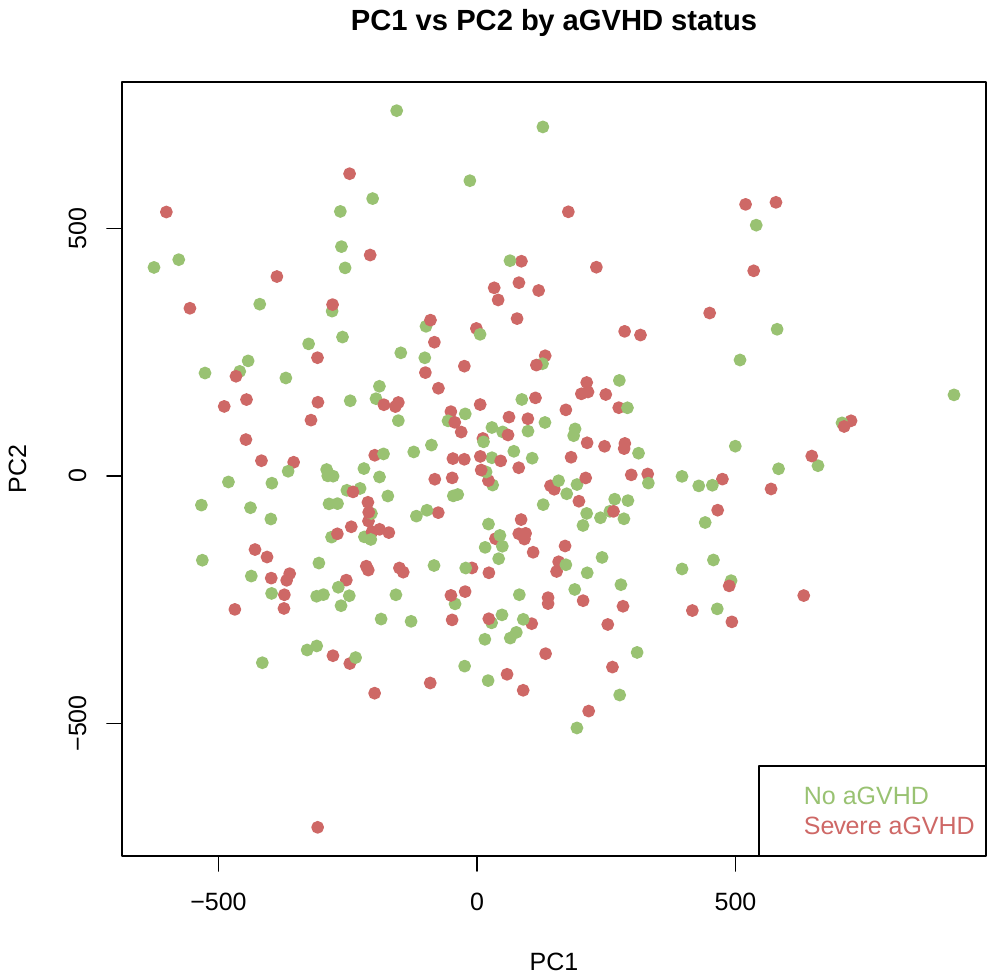

**Supplementary Figure 4: *ROC curve of classifier performance.*** *Plot A shows the performance of the variable probe based (unsupervised approach) classifier which used the top 10,000 most variable CpG sites as input, during internal cross validation on the training dataset. Plot B shows the performance of the variability based classifier on the independent validation cohort, with an AUC of 0.523, a sensitivity of 50.0% and a very poor specificity of 51.4%.*

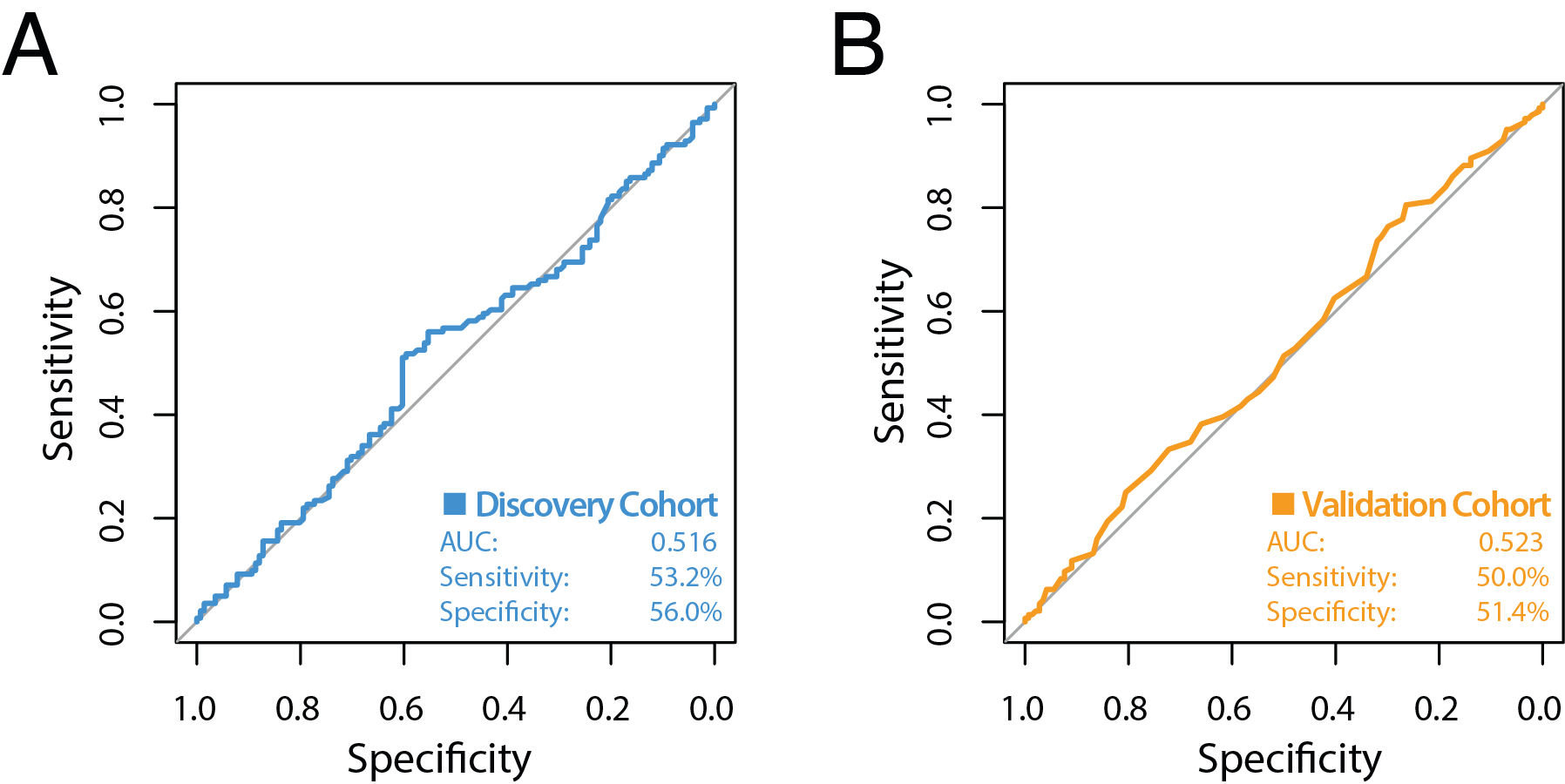

***Supplementary Table 1: Cell composition estimations for discovery and validation cohorts.*** *Immune cell proportions estimated from DNA methylation data using the Houseman reference based method. Discovery and replication cohorts were well matched for predicted cell proportions between no aGVHD and severe aGVHD groups.*

|  | **Discovery study (n=282)** | | | **Validation study (n=288)** | | |
| --- | --- | --- | --- | --- | --- | --- |
| Cell type | No aGVHD (n=141) | Severe aGVHD (n=141) | P-value | No aGVHD (n=144) | Severe aGVHD (n=144) | P-value |
| CD8T | 0.07 | 0.06 | 0.24 | 0.05 | 0.05 | 0.87 |
| CD4T | 0.11 | 0.11 | 0.65 | 0.11 | 0.11 | 0.41 |
| NK | 0.01 | 0.01 | 0.56 | 0.02 | 0.01 | 0.63 |
| B | 0.04 | 0.04 | 0.90 | 0.04 | 0.04 | 0.51 |
| Mono | 0.07 | 0.07 | 0.73 | 0.08 | 0.08 | 0.99 |
| Gran | 0.67 | 0.68 | 0.87 | 0.68 | 0.69 | 0.43 |
